# EndoTime: Non-categorical timing estimates for luteal endometrium

**DOI:** 10.1101/2021.08.02.21261452

**Authors:** Julia Lipecki, Andrew E Mitchell, Joanne Muter, Emma S Lucas, Komal Makwana, Katherine Fishwick, Joshua Odendaal, Amelia Hawkes, Pavle Vrljicak, Jan J Brosens, Sascha Ott

## Abstract

**STUDY QUESTION:** Can the accuracy of timing of luteal phase endometrial biopsies based on urinary ovulation testing be improved by measuring the expression of a small number of genes and a continuous, non-categorical modelling approach?

**SUMMARY ANSWER:** Measuring the expression levels of six genes (*IL2RB, IGFBP1, CXCL14, DPP4, GPX3*, and *SLC15A2*) is sufficient to obtain substantially more accurate timing estimates and assess the reliability of timing estimates for each sample.

**WHAT IS KNOWN ALREADY:** Commercially available endometrial timing approaches based on gene expression require much larger gene sets and use a categorical approach that classifies samples as pre-receptive, receptive, or post-receptive.

**STUDY DESIGN, SIZE, DURATION:** Gene expression was measured by RT-qPCR in 260 endometrial biopsies obtained 4 to 12 days after a self-reported positive home ovulation test. A further 36 endometrial samples were profiled by RT-qPCR as well as RNA-sequencing.

**PARTICIPANTS/MATERIALS, SETTING, METHODS:** A computational procedure, named ‘EndoTime’, was established that models the temporal profile of each gene and estimates the timing of each sample. Iterating these steps, temporal profiles are gradually refined as sample timings are being updated, and confidence in timing estimates is increased. After convergence, the method reports updated timing estimates for each sample while preserving the overall distribution of time points.

**MAIN RESULTS AND THE ROLE OF CHANCE:** The Wilcoxon Rank Sum Test was used to confirm that ordering samples by EndoTime estimates yields sharper temporal expression profiles for held-out genes (not used when determining sample timings) than ordering the same expression values by patient-reported times (*GPX3*: *p* < 0.005; *CXCL14*: *p* < 2.7e-6; *DPP4*: *p* < 3.7e-13). Pearson correlation between EndoTime estimates for the same sample set but based on RT-qPCR or RNA-sequencing data showed high degree of congruency between the two (*p* = 8.6e-10, R^2^ = 0.687).

**LIMITATIONS, REASONS FOR CAUTION:** Timing estimates are predominantly informed by glandular gene expression and will only represent the temporal state of other endometrial cell types if in synchrony with the epithelium. Methods that estimate the day of ovulation are still required as these data are essential inputs in our method. Our approach - in its current iteration – performs batch correction such that larger sample batches impart greater accuracy to timing estimations. In theory, our method requires endometrial samples obtained at different days in the luteal phase. In practice, however, this is not a concern as timings based on urinary ovulation testing are associated with a sufficient level of noise to ensure that a variety of time points will be sampled.

**WIDER IMPLICATIONS OF THE FINDINGS:** Our method is the first to assay the temporal state of luteal-phase endometrial samples on a continuous domain. It is freely available with fully shared data and open source software. EndoTime enables accurate temporal profiling of any gene in luteal endometrial samples for a wide range of research applications and, potentially, clinical use.

**STUDY FUNDING/COMPETING INTEREST(S):** This study was supported by a Wellcome Trust Investigator Award (Grant/Award Number: 212233/Z/18/Z) and the Tommy’s National Miscarriage Research Centre. None of the authors have any competing interests. JL was funded by the Biotechnology and Biological Sciences Research Council (UK) through the Midlands Integrative Biology Training Partnership (MIBTP).

**TRIAL REGISTRATION NUMBER:** N/A.

## Introduction

Menstruation is the defining characteristic of the endometrium in humans and higher primates, a trait otherwise found in only a handful of non-primate species (Bellofiore *et al*., 2017). As a consequence of menstruation, the endometrium undergoes iterative cycles of tissue regeneration, rapid proliferation and differentiation, which cumulate in a transient window of implantation during the midluteal phase of the cycle. The window of implantation represents an inflection point in the cycle, after which the endometrium either breaks down or is transformed into the decidua of pregnancy, a specialized matrix that accommodates the placenta throughout gestation (Gellersen and Brosens, 2014). Endometrial cyclicity is driven by the rise and fall in ovarian estrogen and progesterone production, triggering coordinated spatiotemporal gene expression changes in resident epithelial, stromal and vascular cells (Wang *et al*., 2020). Further, the midluteal window of implantation heralds the start of intense tissue remodelling, characterised not only by abrupt and dramatic changes in epithelial gene expression (Wang *et al*., 2020), differentiation of stromal cells in pre-decidual cells (Lucas *et al*., 2020), and angiogenesis (Demir *et al*., 2010), but also influx of circulating innate immune cells (Strunz *et al*., 2021), most prominently uterine natural killer cells (Brighton *et al*., 2017), as well as non-hematopoietic bone marrow-derived progenitor cells (Diniz-da-Costa *et al*., 2021).

It is widely accepted that pathological cues that interfere with the sequence of endometrial events leading to a functional implantation window causes reproductive failure, including recurrent implantation failure (Koot *et al*., 2016) and recurrent pregnancy loss (Lucas *et al*., 2020). However, it has proven challenging to parse the precise underlying mechanisms. There are multiple challenges intrinsic to endometrial research, including heterogeneity in the cellular composition of endometrial biopsies (Suhorutshenko *et al*., 2018), intrinsic inter-cycle variability in local immune cells (Brighton *et al*., 2017), and - most prominently - the rapid temporal changes in gene expression across the luteal phase (Wang *et al*., 2020). Accurate timing information is therefore critical in endometrial analysis. While the average length of menstrual cycles is 28 days, there is considerable intra- and inter-individual variation (Soumpasis *et al*., 2020). A pragmatic solution is to schedule biopsies relative to pre-ovulatory luteinising hormone (LH) surge (Tewary *et al*., 2020). A prospective study on a small cohort of healthy women (n = 40) reported that the urinary LH surge occurs mostly within one day prior to ovulation, although the range was 4 days (Johnson *et al*., 2015; Roos *et al*., 2015). Further, the rise in urinary pregnanediol-3-glucuronide, a progesterone metabolite, is more variable, occurring over a range of 5 days after ovulation (Johnson *et al*., 2015; Roos *et al*., 2015). Thus, while timing of endometrial biopsies relative to clinical markers of ovulation is useful and convenient, it does not ensure comparable exposures to progesterone stimulation.

A complementary strategy is to infer timing by analysing the endometrial phenotype. Histological dating using the Noyes criteria was the foundational approach (Noyes *et al*., 1950), but its accuracy has been brought into question (Coutifaris *et al*., 2004; Murray *et al*., 2004). Alternative methods for timing are based largely on detection of proteins, transcripts or microRNAs that mark the putative implantation window (Giudice and Saleh, 1995; Lessey, 1998; Develioglu *et al*., 1999; Dubowy *et al*., 2003; Kliman *et al*., 2006; Aghajanova *et al*., 2009; Sha *et al*., 2011; Zhang *et al*., 2012). In addition, two computational approaches for the prediction of the window of implantation are currently available commercially. The ERA (Endometrial Receptivity Analysis) method was initially based on customised microarrays to measure the expression of 238 genes (Díaz-Gimeno *et al*., 2011; Ruiz-Alonso *et al*., 2013). The ER Map/ER Grade approach uses RT-qPCR measurements of 40 genes (Enciso *et al*., 2018). However, both methods time biopsies categorically as either pre-receptive, receptive, or post-receptive, which not only severely limits temporal resolution but also risks misclassification of samples at the boundary of these time windows. At present, there are no cost-effective, validated methods to assess luteal phase endometrium in a continuous, time-dependent domain.

This study describes the development and validation of an expression-based assay that reflects time as a continuous measurement of days and hours, using a discrete set of temporally-sensitive genes. This method, termed EndoTime, is freely available as open source software.

## Materials and Methods

### Ethics

The study was approved by the NHS Research Ethics Committee, Hammersmith and Queen Charlotte’s & Chelsea Research Ethics Committee (1997/5065), and Tommy’s National Reproductive Health Biobank (REC reference: 18/WA/0356). All samples were obtained with written informed consent and in accordance with The Declaration of Helsinki (2000) guidelines.

### Endometrial sample collection

Endometrial biopsies were obtained from women attending the Implantation Clinic, a dedicated research clinic at University Hospitals Coventry and Warwickshire (UHCW) National Health Service Trust. Surplus tissues from endometrial biopsies obtained for diagnostic purposes were used for this study. Participants were instructed to use over-the-counter urinary ovulation tests and contacted the clinic on the day of a positive test or soon after. An endometrial biopsy was then scheduled 4 to 12 days after a positive test. The timing of endometrial biopsies were designated as LH+(day), i.e. the number of days following a positive urinary ovulation test. Following transvaginal ultrasound assessment to exclude overt pelvic pathology, an endometrial biopsy was obtained using a Wallach Endocell™ endometrial sampler. All samples were immediately portioned with one part stored in RNALater Stabilization Solution (Sigma-Aldrich, Dorset, UK), one part snap frozen in liquid nitrogen, and one part fixed in 10 % formalin for immunohistochemistry.

### RT-qPCR

Total RNA was extracted from endometrial biopsies immersed in RNALater Stabilization Solution (Merck, New Jersey, USA) within 1 min of sampling using STAT-60 (AMS Biotechnology, Oxford, UK), according to the manufacturer’s instructions. Reverse transcription was performed from 1µg RNA using the Quantitect Reverse Transcription Kit (QIAGEN, Manchester, UK) and cDNA was diluted to 10 ng/µl equivalent before use in qPCR. Amplification was performed on a Quant5 Real-Time PCR system (Applied Biosystems, Paisley, UK) in 10 µl reactions using 2×QuantiFast SYBR Green PCR Master Mix containing ROX dye (QIAGEN), with 300 nM each of forward and reverse primers. *L19* was used as a reference gene. Primer sequences and information of marker genes are tabulated in Supplementary Table 1.

### RNA-sequencing

RNA was purified using RNA STAT-60 (AMS Bio) according to manufacturer’s instructions and treated using Amplification Grade DNase I (Invitrogen) followed by ethanol precipitation and clean-up. Quality control, library preparation and sequencing were performed by the Wellcome Trust Centre for Human Genetics. Libraries were prepared using the Illumina TruSeq Stranded mRNA sample prep kit according to manufacturer’s instructions. Paired-end 75bp sequencing was performed on Illumina HiSeq4000.

### Imaging

Endometrial biopsies were fixed overnight in 10% neutral buffered formalin at 4°C and wax embedded in Surgipath Formula ‘R’ paraffin using the Shandon Excelsior ES Tissue processor (ThermoFisher). Tissues were sliced into 3 μM sections on a microtome and adhered to coverslips by overnight incubation at 60°C. Deparaffinization, antigen retrieval (pH 9), antibody staining, hematoxylin counter stain and DAB colour development were fully automated in a Leica BondMax autostainer (Leica BioSystems). Tissue sections were stained for CD56 (a uNK-specific cell surface antigen) using a 1:200 dilution of concentrated CD56 antibody (NCL-L-CD56-504, Novocastra, Leica BioSystems). Stained slides were de-hydrated, cleared and cover-slipped in a Tissue-Tek Prisma Automated Slide Stainer, model 6134 (Sakura Flinetek Inc. CA, USA). Bright-field images were obtained on a Mirax Midi slide scanner using a 20× objective lens and opened in Panoramic Viewer v1.15.4 (3DHISTECH Ltd, Budapest, Hungary).

### Pre-processing of qPCR data

RT-qPCR data to be used as input for EndoTime initially had the sign for all ΔCT values inverted in order to positively correlate with gene expression. The data was then assessed for outliers in expression of any of the six genes, leaving 260 endometrial samples. Expression values in individual samples in Data Set I were normalised to a scale of zero to one per gene and then adjusted by a batch-specific additive constant as a modest batch effect correction, making mean expression values equal in each batch. Batch correction was not necessary for samples in Data Set II, as RT-qPCR analysis was performed as a single batch.

### Pre-processing of rLH+ values

EndoTime modelling required that reported sample time points be converted from an ordinal to a continuous domain, a process that was undertaken in two steps. First, the addition of random noise between -0.5 and 0.5 sampled from a uniform distribution to each reported LH+ (rLH+) value. Second, samples were sorted in ascending order according to these updated timings, and the timings were smoothed using linear regression. This procedure allowed for each sample to be spaced evenly throughout the defined time course in a non-discrete manner, but close to its original rLH+ value, an approach that was considered robust in the presence of samples with unusually high or low reported timing values.

### EndoTime Method

The approach for modelling via EndoTime relies upon an iteration of temporal gene expression profile refinement followed by the application of these refined profiles to estimate sample timings. Continuous rLH+ values generated during pre-processing were used to form initial expression profiles specific to each panel gene, which were then partitioned into windows of equal size (Supplementary Fig. S1). A normal distribution was used to model gene expression inside the time window with a weighted mean based on the relative distance of points from the mean and standard deviation inferred from samples inside the window (Supplementary Fig. S1 and S2). Each window represented a singular time point derived from the median of binned reported time points. The first iteration utilised a bin size of 80 samples, which decreased by 10 with each successive iteration to a minimum of 20.

Each sample within the data set was then assessed individually for its likely timing. The expression values for each marker gene within each window were used to generate a probability density curve based on the likelihood that the expression values observed in a sample were drawn from the distribution seen in the windowed data. This resulted in a set of six probability density curves being generated per sample, with each curve representing the results of attempting to estimate timing for the sample based on one gene alone, with associated curve maxima suggesting the time point with the greatest likelihood. Utilising a shrinking bin size allowed for the first iteration of the process to filter out the majority of noise introduced by the unreliable rLH+ into the data while subsequent iterations refined estimations, while enforcing a minimum bin size ensured that the curves were smooth and presented a single clear maximum.

This process of generating six individual probability density curves also allowed for an assessment to be made regarding the congruency of their peak maxima and therefore the consistency with which each gene provided the same timing estimate. Synchronous samples were those wherein the six maxima all suggested a similar estimate, while asynchronous samples were those that presented conflicting estimations; an ‘asynchrony’ score was provided to each sample based on the standard deviation among all six maxima, which describes how coherently the aforementioned probability density curve-based approach provides a singular timing estimate.

Consolidating these six curves into a single aggregate curve allowed for the identification of a maximum in a single curve, which was used as the new time point estimation for the sample. A window-based approach was used when consolidating the six individual curves into one, with bin sizes equal to those used when generating the underlying reference profiles of panel gene expression. This process iterated until convergence, with each iteration undertaking both refinement of temporal profiles and time point estimation. The difference between the estimations provided by the preceding and current iteration was measured using the Euclidean distance and convergence was declared once the distance falls below 2.

During the modelling process, the absolute values of sample timings could deviate from the desired range as our method was primarily geared to optimising the correct order of samples, rather than retaining the original unit associated with timings. To ensure that EndoTime outputs are in line with original units, the raw timings obtained by modelling were converted following the last iteration such that the overall distribution of patient-reported LH times is approximately matched by the EndoTime output.

### Pre-processing of RNA-seq Data

RNA-seq libraries were mapped to the hg19 human genome assembly (2014) using Bowtie (version no. 2.2.3), TopHat (version no. 2.0.12) and Samtools (version no. 0.1.19) and reads mapped to features were counted via HTSeq (version no. 0.6.1) prior to Transcripts Per Million (TPM) normalisation.

In order to apply EndoTime to RNA-seq data, an approach was developed to convert read counts of EndoTime panel genes to pseudo-RT-qPCR data. Transcripts Per Million for each of the six timing panel genes were initially log2-transformed and then transformed to match the mean and standard deviation for each respective gene in the RT-qPCR data of Data Set I, with all processing performed in R (version no. 4.0.2) using base functions.

### Statistical Analyses

To assess the improvement in timing accuracy, we used a cross validation approach. Sample timings were estimated using EndoTime with a panel of only five genes, holding out the expression data for one gene. Expression data for the held-out gene was ordered a) by patient-reported times (denoting this vector as *v*_*P*_), b) by EndoTime timing estimates (denoted as *v*_*E*_), and c) by expression level, in ascending order if the expression level of the held-out gene increases over time and descending order otherwise (denoted as *v*_*G*_). As patient-reported times are integer values with a unit of days, breaking ties needed to be resolved in order to compare directly against the other vectors. This was done by ordering samples of the same day in ascending or descending order according to the expression level of the held-out gene. We applied the Wilcoxon rank sum test to check whether the absolute values of the differences *v*_*G*_ *- v*_*P*_ were greater than for *v*_*G*_ *- v*_*E*_ in a single-sided test. A significant p-value indicates that the order of samples provided by EndoTime is closer to the perfect order. In this setting, breaking the ties for patient-reported times as described above yields the largest p-value among all possible resolutions of ties, meaning that statistical significance may be under-stated but not over-stated with this approach as the p-value computed is an upper bound for the p-value that could be obtained if patient-reported times were more finely resolved. This process was repeated six times, holding out one panel gene at a time, and p-values Bonferroni-corrected for multiple testing.

RNA-seq data was examined via Principal Component Analysis in MATLAB following transformation of raw counts using the rlog function from the R library DESeq2 (version no. 1.30.1).

### Data and Software Availability

The EndoTime software and software documentation are available on GitHub at https://github.com/AE-Mitchell/EndoTime under the GNU General Public License Version 3 (29 June 2007). The RT-qPCR data files are included in the GitHub repository as is the code used for pre-processing of RT-qPCR data. The RNA-seq data are available on GEO (accession GSE180485).

## Results

The EndoTime method was developed using two sample sets of luteal phase endometrial biopsies. Demographic information for both sample sets are presented in Supplementary Table 2. Sample set 1 consisted of 260 endometrial biopsies assayed by RT-qPCR in nine batches. Out of the 260 biopsies, 98 were obtained from women with a history of recurrent miscarriage (defined here as 3 or more consecutive pregnancy losses), 81 from women with repeated IVF failure (i.e. no positive pregnancy test following 3 or more transfers of day 5 blastocysts) and the remaining 81 biopsies from control subjects. Sample set 2 consisted of 36 endometrial biopsies assayed by RT-qPCR and RNA-seq as a single batch. The distribution of endometrial biopsies relative to the patient-reported positive urinary ovulation test in both sample sets is shown in Supplementary Table 3.

### The EndoTime Method

Timing estimates of endometrial biopsies should ideally rely on temporal reference profiles of marker genes that span the entire luteal phase and are free of noise, as illustrated by synthetic data in Figure 1A. In reality, only a limited number of biopsies can be sampled (Fig. 1B), and patient-reported days since a positive urinary ovulation test (rLH+) will be subject to a degree of error and noise as simulated in Figure 1C, thus obscuring the true temporal expression patterns. We observed that the simulated data show a very similar pattern to real-world data (Fig. 1D), illustrating the practical relevance of this theoretical framework. EndoTime aims to minimise the impact of this source of noise by recovering the original expression patterns and thereby allowing for more accurate estimation of endometrial timing.

**Figure 1:**
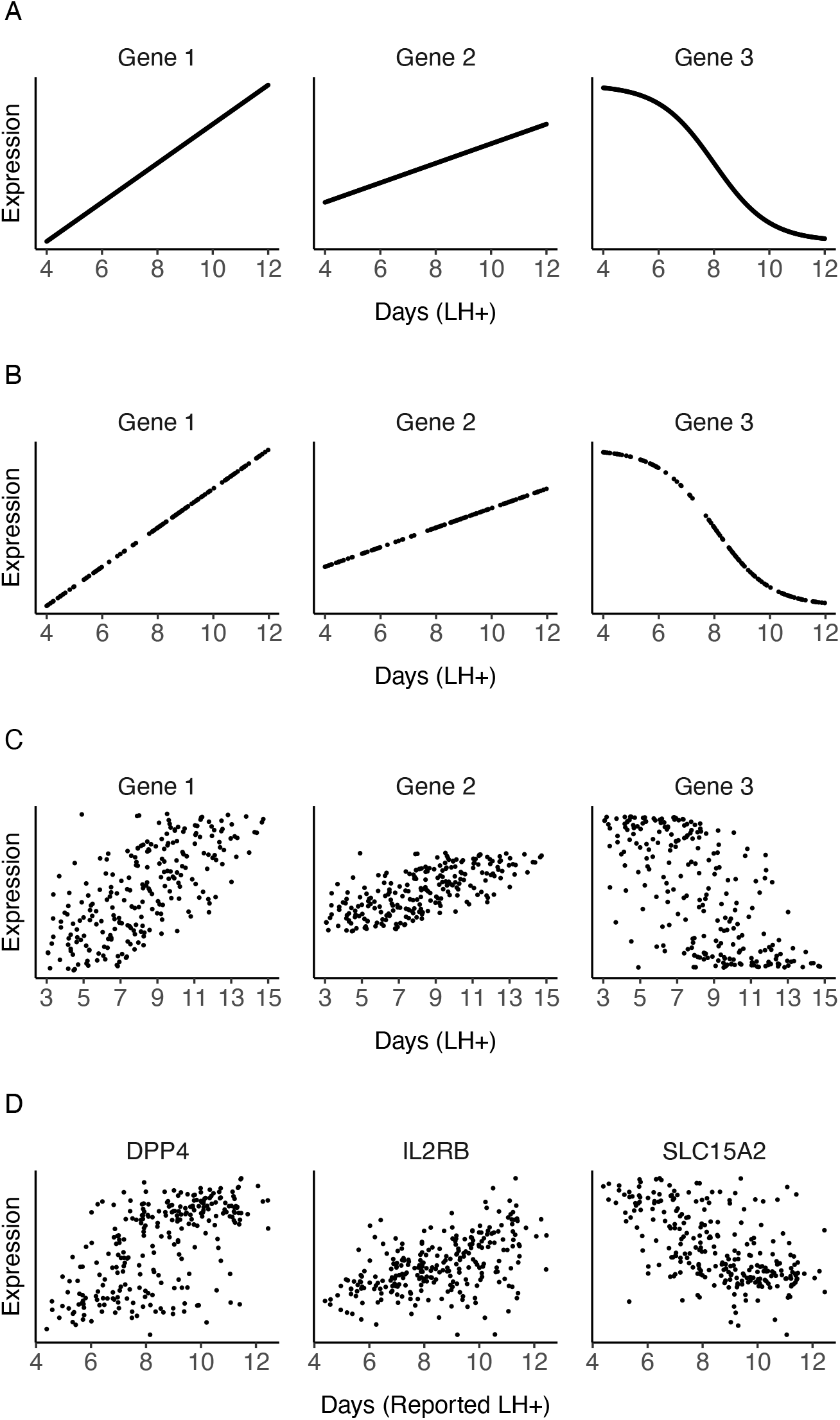
Effect of sampling and noise on measured temporal profiles. (A) Ideal expression curves for three artificial genes with infinite sampling density and without noise. (B) Simulated data as in A but with limited uniform sampling over the time axis more reflective of real-world biopsy availability. (C) Data simulated as in B with random noise added to time points (noise sampled from the normal distribution, mean = 0, standard deviation = 2) to reflect uncertainty in reporting. (D) Expression measurements of three genes in clinical samples with patient-reported timings. The observed gene expression patterns are a good match for anticipated patterns simulated in C in terms of noise level and fuzzy appearance of temporal profiles.

Accomplishing this goal requires us to solve a Chicken and Egg problem: inferring the correct time point for a given biopsy requires accurate reference expression profiles, but recovery of these profiles relies on accurately timed biopsies. Our solution is to apply an iterative approach, using the initial rLH+ time points to model expression profiles while accounting for uncertainty (Fig. 2A), then updating biopsy timings for all samples based on the modelled reference profiles (Fig. 2B). These two steps are iterated, with reference profiles gradually becoming less noisy as timing estimates are improved in a stepwise manner (Fig. 2C). The process is repeated until convergence, defined as a minimal overall change of sample timings from one iteration to the next.

**Figure 2:**
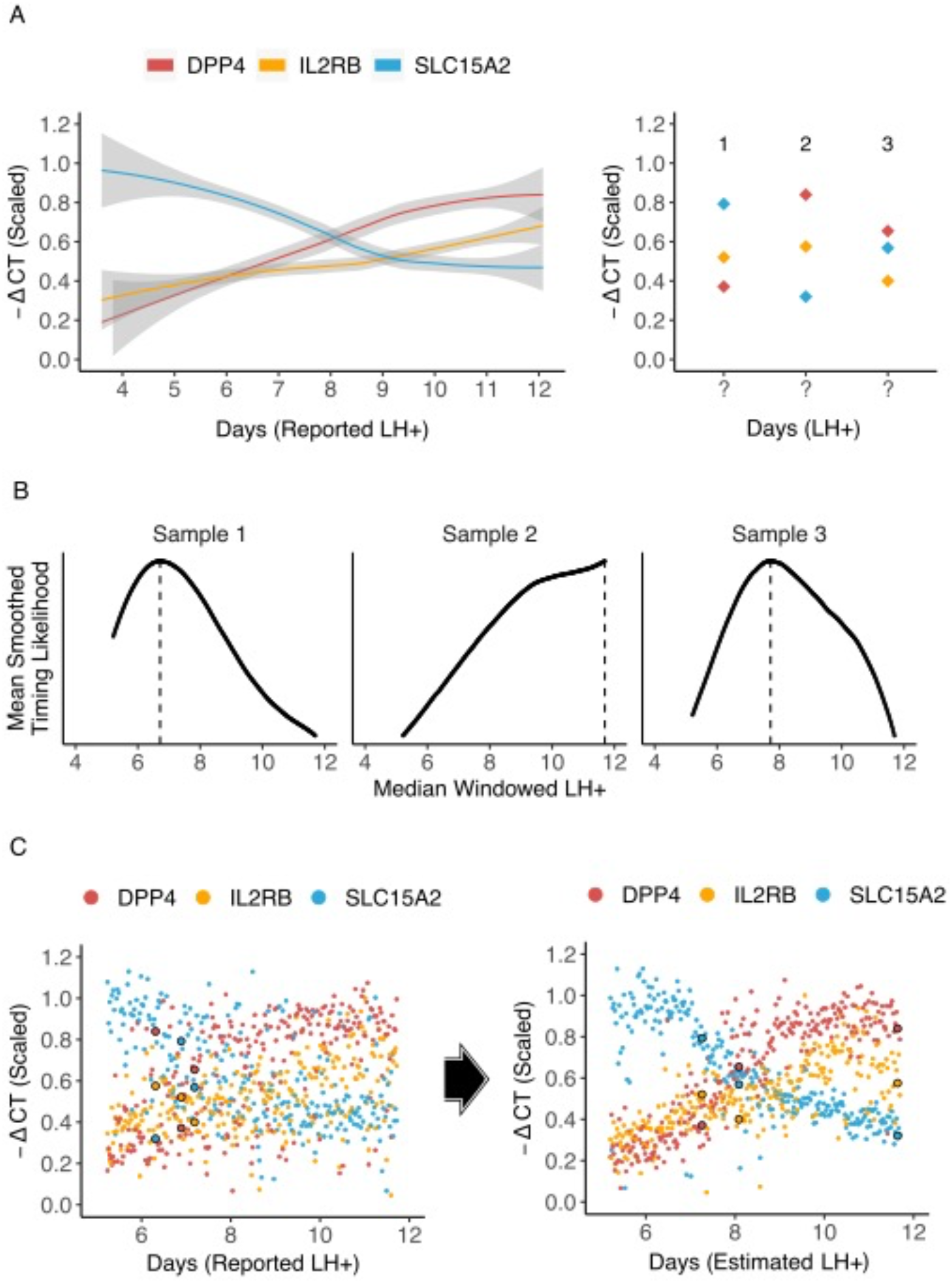
Illustration of one iteration of the EndoTime modelling process. (A) Computing temporal profiles. Left: Regression curves fit to expression data for three genes of the timing marker panel. Right: Expression values for three samples from the training data are the basis for re-evaluating timings of these samples. (B) Temporal profiles learned in A are used to improve time point estimates. For each sample, the likelihood of each time point is computed, with suggested sample timing represented by peak maxima for each sample. (C) Improved time point estimates provide improved temporal profiles. Left: Expression data arranged according to patient-reported LH+. Values for the three samples from A and B circled. Right: Expression data re-arranged according to new time estimates obtained in B. Expression curves are visibly tighter and more distinct after just one iteration. EndoTime repeats this process until convergence.

Modelling temporal expression profiles is done using a window-based approach that considers samples in individual segments of the time domain and modelling their mean and variance as a normal distribution (Supplementary Fig. 1). The size of the windows is gradually decreased from iteration to iteration as sharper temporal profiles allow for a more detailed model of the reference profiles. The position of samples inside a window is taken into account when computing the means such that samples near the centre of the window have stronger influence than samples near the edge (Supplementary Fig. 2).

The modelled temporal profiles (Fig. 2A) are then used to compute probability density functions for each sample and each marker gene, which show how likely each time point is for the given sample as judged by the reference profile of a single marker gene. Joint probability density functions are then computed, showing likelihood of sample timing on the basis of the reference profiles for all marker genes (Fig. 2B). The maxima of these functions are then identified for each sample, which provide maximum likelihood estimates for the most appropriate sample timing. The iterated process of updating the reference profiles and updating sample timings gradually refines the reference profiles and increases the certainty in timing estimates (Fig. 2C, Supplementary Fig. S3).

### Validation of EndoTime Method

We applied the EndoTime method on sample set 1, comprising of 260 luteal endometrial samples. We used a leave-one-out cross-validation approach for the set of marker genes used, inferring timings based on five genes while not using the expression data of the held-out, sixth gene. We hypothesized that EndoTime estimates will yield sharper, less noisy temporal profiles for temporally regulated genes. If samples were ordered merely to fit the data of five genes without inferring the true order of samples, then the temporal profile for the held-out gene would not improve. This process was repeated six times, holding out one gene at a time. We found that the temporal profile of each held-out gene became tighter after EndoTime with expression values deviating less from the temporal trajectory when compared to profiles plotted using patient-reported times (Fig. 3, right and left panels, respectively). This effect was particularly pronounced for *CXCL14, DPP4*, and *GPX3*. The Wilcoxon Rank Sum Test was used to confirm that the improvement in temporal expression profiles for three held-out genes was statistically significant (see Methods; *GPX3*: p < 0.005; *CXCL14*: p < 2.7e-6; *DPP4*: p < 3.7e-13). The other genes, though visually appearing tighter, did not test significantly under the conservative testing approach used here which resolves breaking ties for patient-reported times in a way that maximises the *p*-value (see Methods). These genes may also be less tightly regulated in the luteal phase. We concluded that EndoTime arranged samples on the time axis in a biologically more accurate manner than patient-reported times.

**Figure 3:**
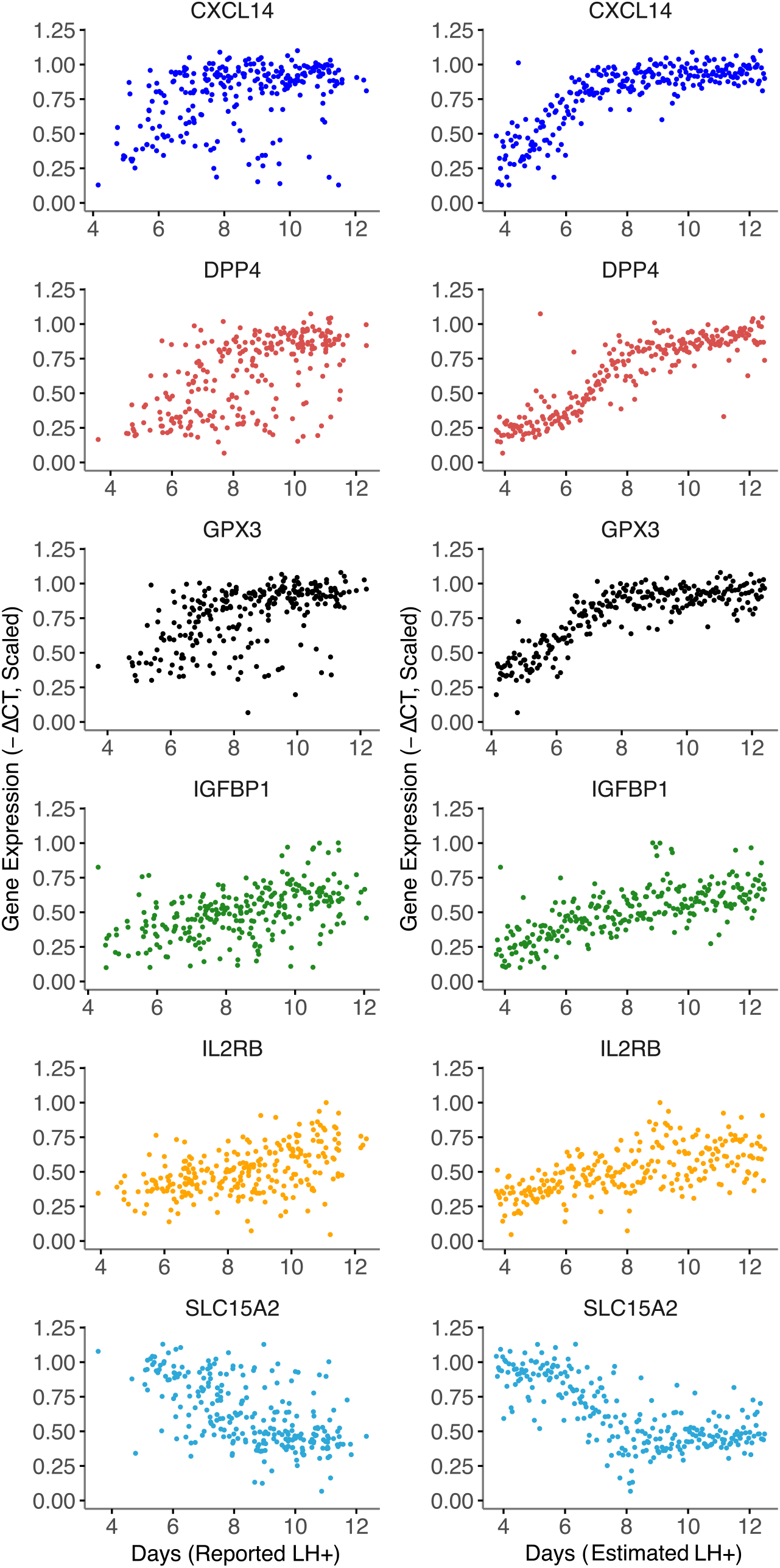
Method validation by leave-one-out approach. Left: Expression measurements for panel genes plotted using time points reported by patients. As original data is only resolved to full days, samples have been moved randomly on the time axis with an average displacement of 6 hours (maximum of 12 hours) to make data visualisation more comparable with EndoTime estimates on continuous domain. Right: temporal profile of each gene after using data of the other five genes to obtain timing estimates for all samples. Substantially sharper profiles show that EndoTime reveals the true order of samples more accurately than clinical records.

### Detecting Asynchronous Samples

While the order of samples computed by EndoTime reduces the variability in temporal profiles substantially, some individual samples appear to be outliers. We queried if it was possible to assess the reliability of estimates on a per-sample basis to enable the automatic detection of the least reliable samples. As EndoTime computes probability distributions of timing for each marker gene individually before aggregating these, the procedure could be compared to a voting scheme where each marker gene has one vote, enabling an assessment of consistency among marker genes. We formulated a score to measure asynchrony between timing estimates based on individual marker genes (Fig. 4A). Samples with a high asynchrony score show large discrepancies between marker genes and account for the most outlying samples (Fig. 4B; Fig. 4C, right panel; and Fig. 4D, bottom panel). By contrast, synchronous samples show consistency among marker genes (Fig. 4C, left panel; Fig. 4D, top panel), and a good fit to the temporal profile (Fig. 4B). We concluded that EndoTime’s asynchrony score can automatically inform the user about unreliable estimates, which may either be due to noise in experimental measurement for the affected samples or reflect asynchronous gene expression in the tissue. The user may decide to remove such samples from the analysis and refine further the temporal profiles and timing estimates for the remaining samples or, alternatively, repeat the cDNA conversion and RT-qPCR assay of samples deemed asynchronous.

**Figure 4:**
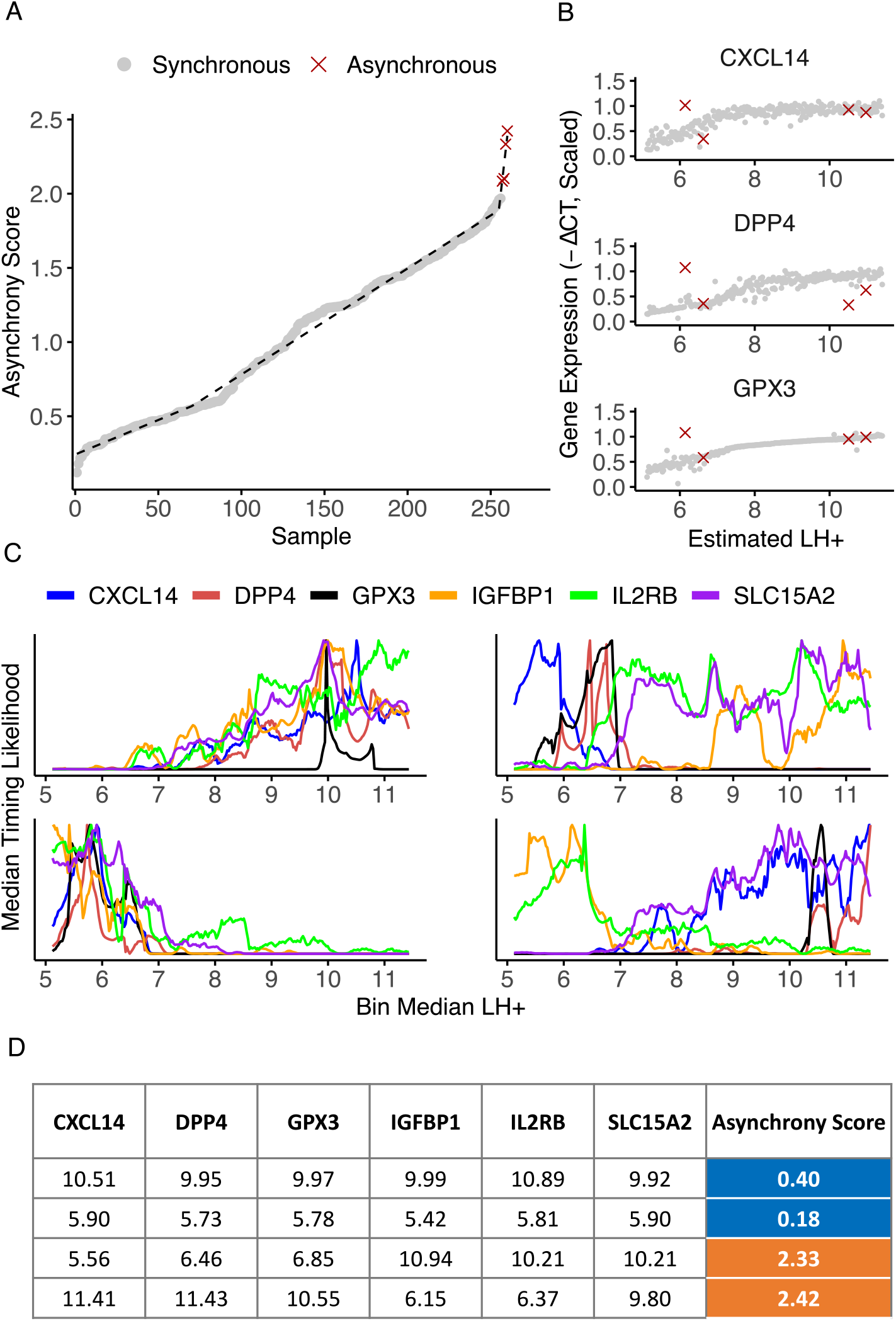
Quantification of sample asynchrony based on consistency between panel genes. (A) Samples are ranked according to their asynchrony score. Breakpoint of segmented linear model designates cut-off point for outliers. (B) Gene expression profiles for three timing panel genes following modelling, with outlying asynchronous samples highlighted. Each sample deemed asynchronous shows discrepant expression values for at least one gene. (C) Timing likelihood for all panel genes for two synchronous samples (left, top and bottom) and two asynchronous samples (right, top and bottom). Synchronous samples exhibit curves with maxima conforming towards a singular predicted time point, while asynchronous samples exhibit contradictory maxima. (D) Time point estimates based on maxima for each panel gene for samples shown in C.

### EndoTime Can be Applied to RNA-seq Data

Using EndoTime analysis of the 260 biopsies in sample set 1 yielded refined gene expression profiles, arranged according to the estimated timings. These profiles can subsequently be utilised alongside new sample sets, an application that is particularly useful if these new sets are not large enough to obtain detailed reference profiles, or if patient-reported timings are not available, which are necessary to initiate the training process.

Sample set 2 consisted of 36 endometrial biopsies for which RT-qPCR as well as RNA-seq data across 33,329 genes was obtained. We used this set to assess if estimates derived from RT-qPCR data would yield comparable results when EndoTime is applied to measurements of the same six marker genes by RNA-sequencing. This necessitated normalising the RNA-seq read counts to make these comparable to the normalised RT-qPCR values in terms of means and variances. As reference profiles were fixed by the modelling exercise for both data types, EndoTime was applied only to carry out a single estimation step for sample timings without updating the temporal profiles. Figure 5 demonstrates that RT-qPCR and RNA-seq time estimates are highly correlated (*p* = 8.6e-10, R^2^ = 0.687). We concluded that meaningful EndoTime estimates can be obtained from RNA-seq data even if there is not enough data to re-train reference profiles.

**Figure 5:**
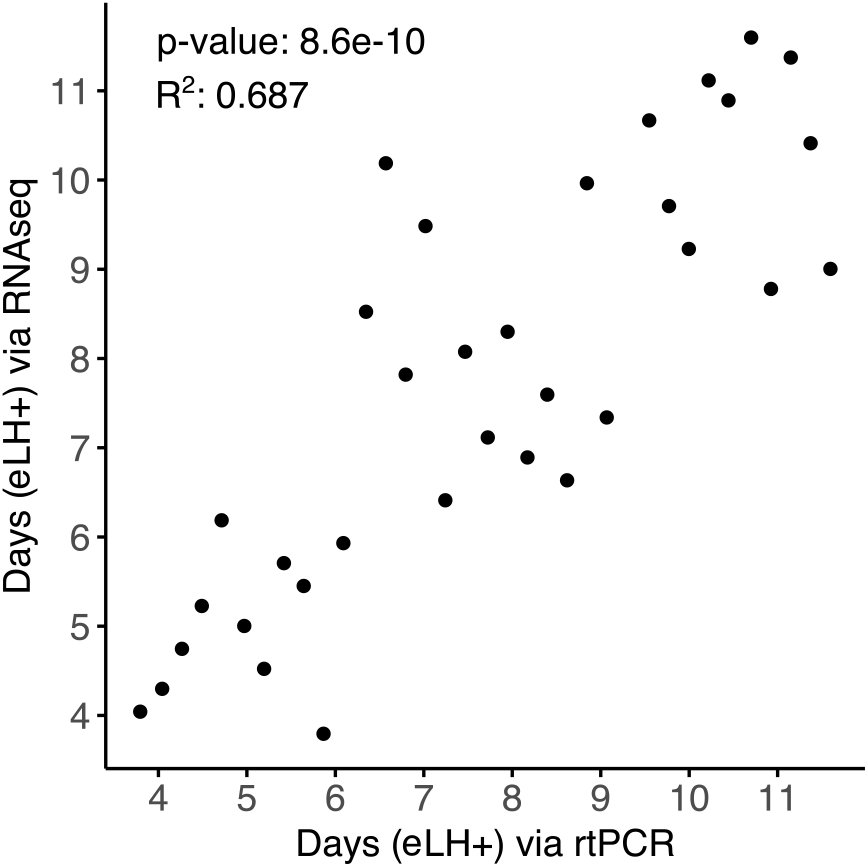
Correlation of predicted time points from qRT-PCR data vs predictions from RNA-seq data of six panel genes.

### Inaccuracy of Reported LH+ Times

A fundamental concern towards reliance upon patient-reported timings provided with clinical biopsies is the potential for inaccuracy. The endometrium is intrinsically dynamic and mistimed samples could confound the diagnosis of underlying pathologies. Histological approaches can provide insights into biopsy timing but require additional processing of samples and appropriate expertise.

EndoTime analysis of the 260 biopsies in sample set 1 revealed a mean difference between reported and estimated LH timing of 1.29 days, with 48 samples showing an estimated deviance of more than two days and 19 samples a deviance of more than three days. One biopsy was estimated 6.22 days later than the rLH+ value. The likelihood of mistiming appeared to be broadly independent of the temporal state of the tissue (Fig. 6A), with deviations occurring throughout the luteal phase. This disparity was also seen upon comparison of patient-reported timings with histological analysis of the tissue samples, the latter of which were congruent with the predictions provided by EndoTime (Fig. 6B).

**Figure 6:**
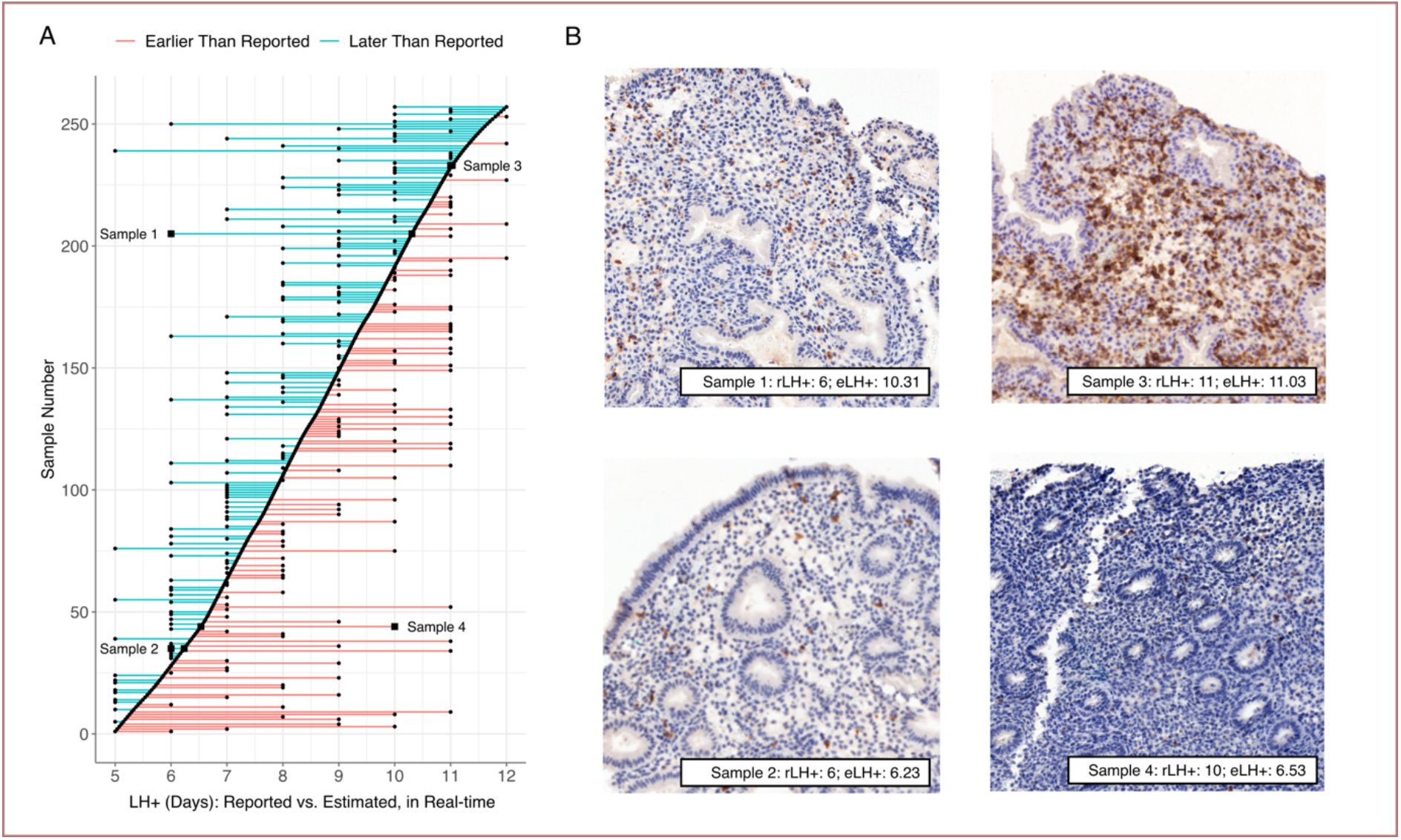
Identification of mistimed samples. (A) Samples shown in order identified by EndoTime (y-axis). EndoTime times shown as smooth curve. Deviations of reported timings from EndoTime timings shown as coloured horizontal lines. (B) Bright-field imaging with staining by the uNK marker CD56 for four samples. These images can be used to verify the progress of tissue development as earlier time points are associated with simple and tubular glands, while corkscrew-shaped glands are associated with biopsies donated during later time points. Samples 1 and 2 appear to be early samples while Sample 3 and 4 are late samples. EndoTime estimates are consistent with this. Reported timings agree for Sample 2 and 3 but are discrepant for Sample 1 and 4 by about four days.

### EndoTime Captures Primary Source of Transcriptomic Variability in Endometrium

Appraisal of the influence of time on transcriptomic variability in comparison to other potential sources of variation, such as interpatient variability, was achieved by performing Principal Component Analysis on the RNA-seq data in sample set 2. The two principal components that explained the largest percentage of variance overlaid RT-qPCR-based EndoTime estimations (Fig. 7), implying that at least 44.1% of variance among 33,329 genes measured can be explained by temporal fluctuations as measured accurately using just the six genes in the EndoTime panel. We conclude that EndoTime captures the primary parameter underlying transcriptomic variability in endometrial biopsies obtained during the luteal phase of the menstrual cycle.

**Figure 7:**
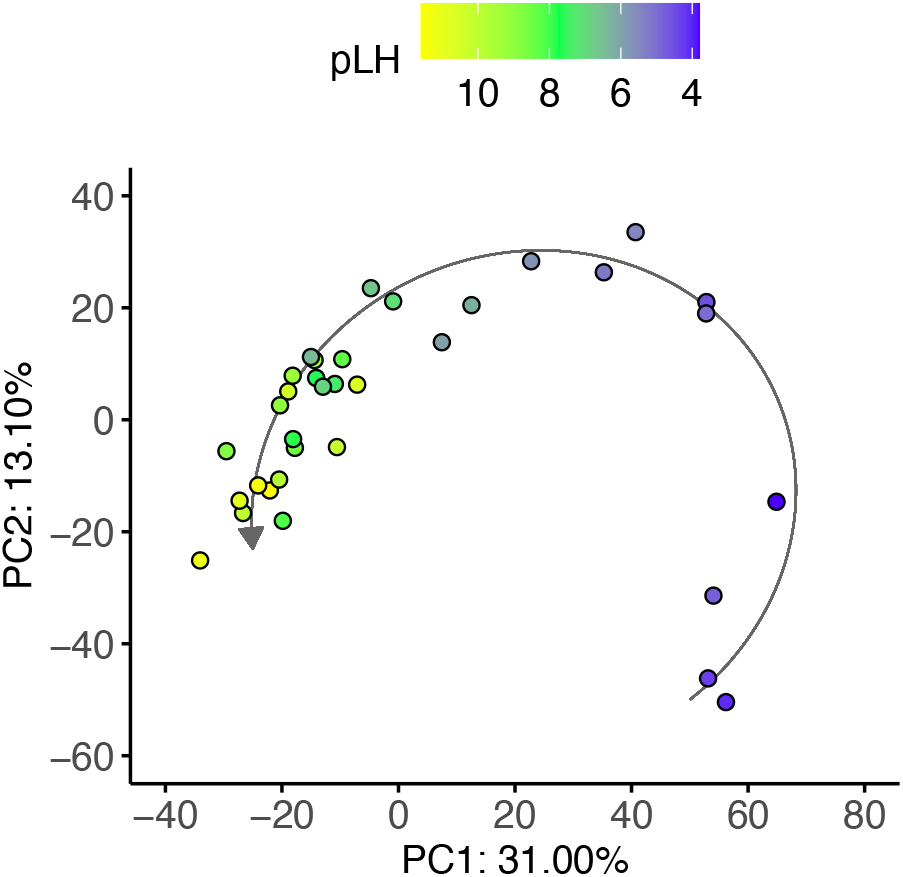
EndoTime estimates capture largest single source of variability in endometrial transcriptomes. PCA performed on 33,329-dimensional RNA-seq data. Colours indicate EndoTime timings inferred from just six genes which are consistent with sample positions in PC 1 and 2 which capture 44.1 percent of transcriptomic variation.

## Discussion

EndoTime utilises the transcriptomic profiles of an informative panel of genes in order to obtain temporal estimates in a continuous domain, rather than making a categorical classification. This avoids misclassifications that are likely when samples are near the temporal threshold between different categorical phases of the cycle and increases resolution of temporal analyses. Given the two existing, transcriptomics-based methods are using only three categories, a misclassification into the neighbouring category implies substantially altered biological interpretation. We have shown the accuracy of EndoTime by leave-one-out validation, which involves removing one panel gene at a time and assessing the sharpening of the temporal profile of the held-out gene. In all cases, the results were comparable to those when using the entire panel, with only minimal increased noise in the estimated timings.

As the measurement of only six genes is required and the software is freely available, EndoTime minimises the obstacles for wide adoption. EndoTime enables any measurements obtained from endometrial biopsies to be interpreted in relation to precise sample timing, thereby revealing the true temporal patterns much more accurately. Importantly, the model training is part of the EndoTime software, enabling the application of EndoTime in other settings, for example with modified sets of panel genes or in different patient cohorts. In fact, EndoTime may be applicable to other tissues and other biological processes if the panel genes are chosen accordingly. We believe that EndoTime has a range of applications in research and may become useful for clinical application as well.

EndoTime provides a good degree of transparency to the user, with each panel gene contributing its own estimate of sample timing, which are then aggregated in a single final time estimate. Estimates based on individual genes that appear inconsistent are reported to the user as asynchrony between panel genes, providing a measure of reliability and highlighting estimates with low confidence. Transcriptomic measurements in a biopsy sample can be plotted against the normal temporal profiles identified by EndoTime, providing a direct visual representation of the evidence for synchrony or asynchrony. Asynchrony could occur as a result of technical errors or biological causes. Although there was no observable correlation between timing errors and any of the three clinical groups that comprise the sample set 1, future work could further investigate correlations of asynchrony with reproductive pathologies in larger data sets.

EndoTime is able to provide timing estimations of greater accuracy as the size of contributing batches increases due to improved batch effect correction in the underlying transcriptomic data used for modelling. Samples in this study were exclusively obtained between 2 and 6 pm, which limits the degree to which timing estimations might be influenced by fluctuations imposed by the circadian clock, such as those associated with *PER2* (Uchikawa *et al*., 2011; Muter *et al*., 2015). EndoTime’s accuracy might be improved via addition of an endometrial circadian gene to the panel and subsequent adjustments to the model should allow for greater timing resolution that takes into account these daily rhythms. Of the six marker genes utilised, four are notably associated with the epithelium, implying that EndoTime estimates are mostly informed by the epithelial compartment of the endometrium.

By transforming RNA-seq measurements to match the distribution of RT-qPCR data prior to modelling via EndoTime, estimates can be obtained that are highly congruent. This conclusion was further supported upon projecting estimated timings over the principal component analysis of RNA-seq data, showing that over 44% of transcriptomic variance between samples can be explained as temporal fluctuations in gene expression. This offers the possibility of applying EndoTime to the transformation and timing estimation of endometrial RNA-seq data. This should broaden the application of EndoTime and inform about temporally sensitive genes which might further improve the gene panel in the future. This may also provide a foundation for dissecting normal temporal changes from changes related to patient cohorts. In addition, it creates potential for developing methods for adjusting the timing of RNA-seq data sets computationally to make these more comparable across patient cohorts.

In summary, EndoTime is a novel open access software which advances the process of timing luteal phase endometrial biopsies along a continuous scale, presenting opportunities for further improvements in terms of its generalisation across the entire endometrium. Its application to a wider range of transcriptomic measurements and in its timing resolution presents potentially far-reaching research and clinical applications.

## Authors’ Roles

JL and AEM contributed to study design, carried out method development, software development, and data analyses. AEM wrote the manuscript and the software documentation. JL contributed to manuscript writing. ESL, JM, KM, KF, JO, and AH processed patient samples and generated experimental data. ESL, JM, and KM provided critical discussions. PV analysed the RNA-seq data, tested the software, and provided critical discussions. JJB obtained patient samples. JJB and SO contributed to study design and writing of the manuscript, provided critical discussions, and provided supervision.

## Acknowledgements

We are grateful to all the women who participated in this research. We also acknowledge the invaluable contribution of Professor Siobhan Quenby and the staff of the Biomedical Research Unit and Tommy’s National Reproductive Health Biobank at University Hospitals Coventry and Warwickshire NHS Trust for facilitating sample collection and processing.

---

**Supplementary Figure 1:**
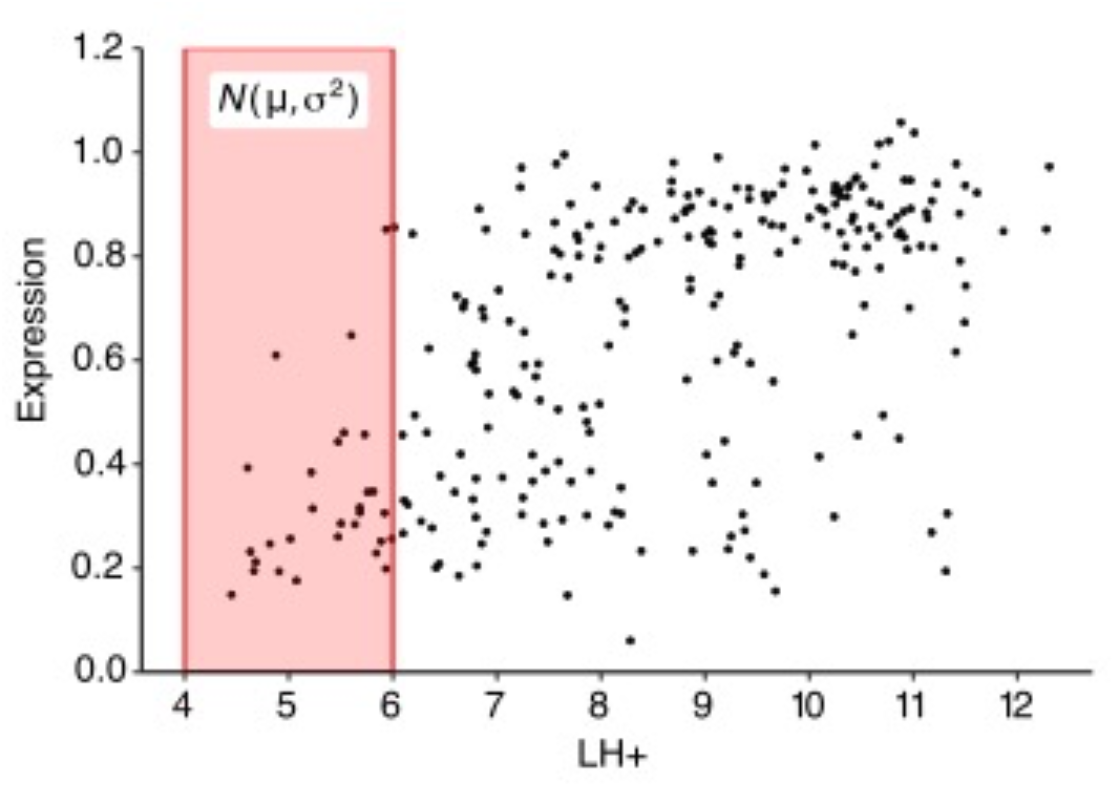
Moving window estimates of expression distribution. Samples are ordered based on current estimates of endometrial timing. Expression at each time point is modelled as a normal distribution. A window (red shading) is placed around a given time point, mean and variance are estimated using only expression levels of samples inside the window. This is repeated for all window positions along the time axis.

**Supplementary Figure 2:**
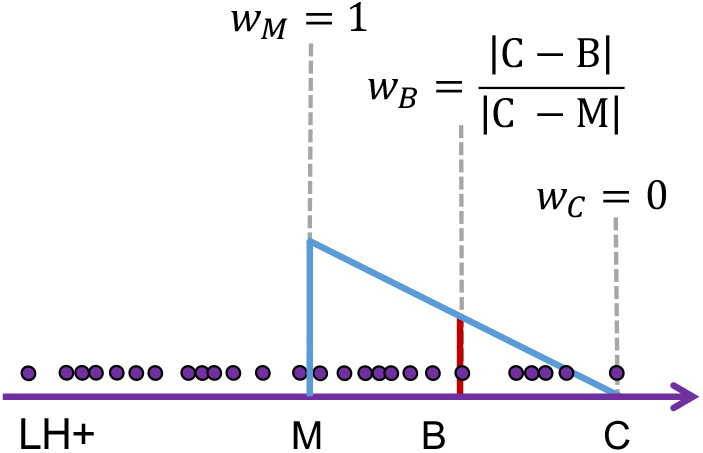
Weighted estimate of means. X axis represents a single window, time estimates for samples inside the window indicated by circles. Each sample contributes to the mean in dependence of proximity to the middle of the window, with closer samples contributing more. M: middle position of the window (weight of 1), C: right-hand-side end of window (weight of 0), B: position of one sample, computation of weight for this sample is illustrated (weight between 0 and 1).

**Supplementary Figure 3:**
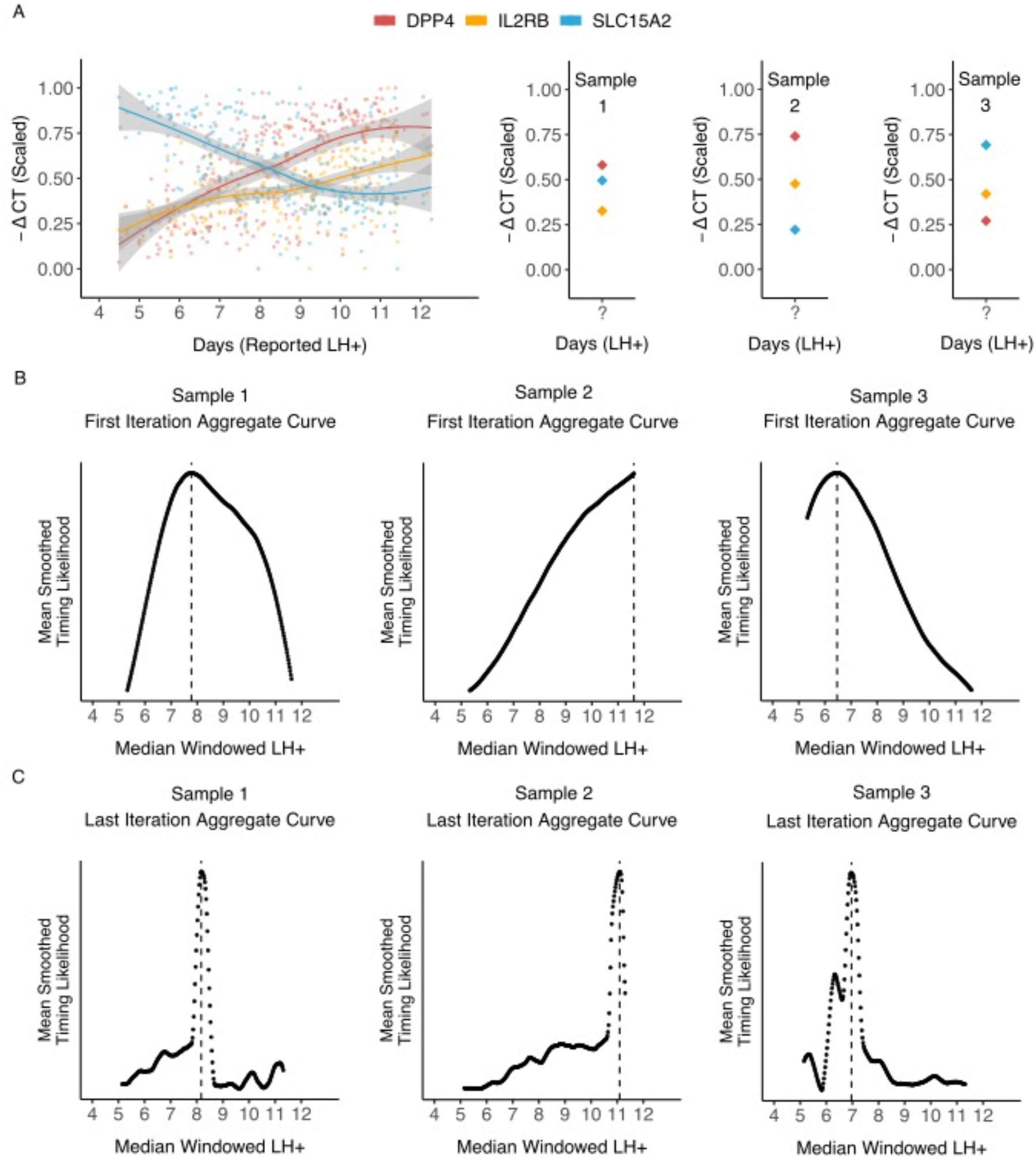
Confidence in estimates increases over multiple iterations. (A) Computing temporal profiles. Left: Regression curves fit to expression data for three genes of the timing marker panel. Right: Expression values for three samples are the basis for re-evaluating timings of these samples. (B) The results of the first iteration of timing estimation for these samples. Each panel gene is used to compute a likelihood that a sample was taken at a particular time point; these are then aggregated into a single likelihood curve per sample, with the peak maxima selected as the temporal estimation for this iteration. (C) The results of the final iteration of timing estimation for each selected sample. Modelling has reached a threshold whereby continued iterations are considered to be of negligible benefit; likelihood curve peaks have sharpened substantially over preceding iterations, providing greater confidence in the estimated time value per sample.

**Supplementary Table 1:** Patient demographics for Sample Set I (qPCR, 260 samples) and II (qPCR & RNA-seq, 36 samples).

|  | Data Set I | Data Set II |
| --- | --- | --- |
| Age [median years ($Q_1 - Q_3$)] | 36 (33-38) | 33 (30-36) |
| BMI [median ($Q_1 - Q_3$)] | 24 (22-27) | 24 (23-27) |
| LH+ day [median ($Q_1 - Q_3$)] | 9 (7-10) | 8 (7-8) |
| First trimester loss [median (range)] | 1 (0-18) | 4 (0-9) |
| Live births [mean (range)] | 0 (0-2) | 0 (0-1) |

**Supplementary Table 2:** Distribution of patient-reported timings in sample sets used in this study.

|  | Sample Timing (rLH+) |  |  |  |  |  |  |  |  |
| --- | --- | --- | --- | --- | --- | --- | --- | --- | --- |
|  | 4 | 5 | 6 | 7 | 8 | 9 | 10 | 11 | 12 |
| Sample Set I: qPCR Samples | 1 | 13 | 29 | 42 | 43 | 44 | 42 | 41 | 5 |
| Sample Set II: qPCR & RNA-seq Samples | 0 | 0 | 3 | 11 | 16 | 6 | 0 | 0 | 0 |

rLH+, reported days since luteal hormone surge, as provided via urinary ovulation testing.

**Supplementary Table 3:**
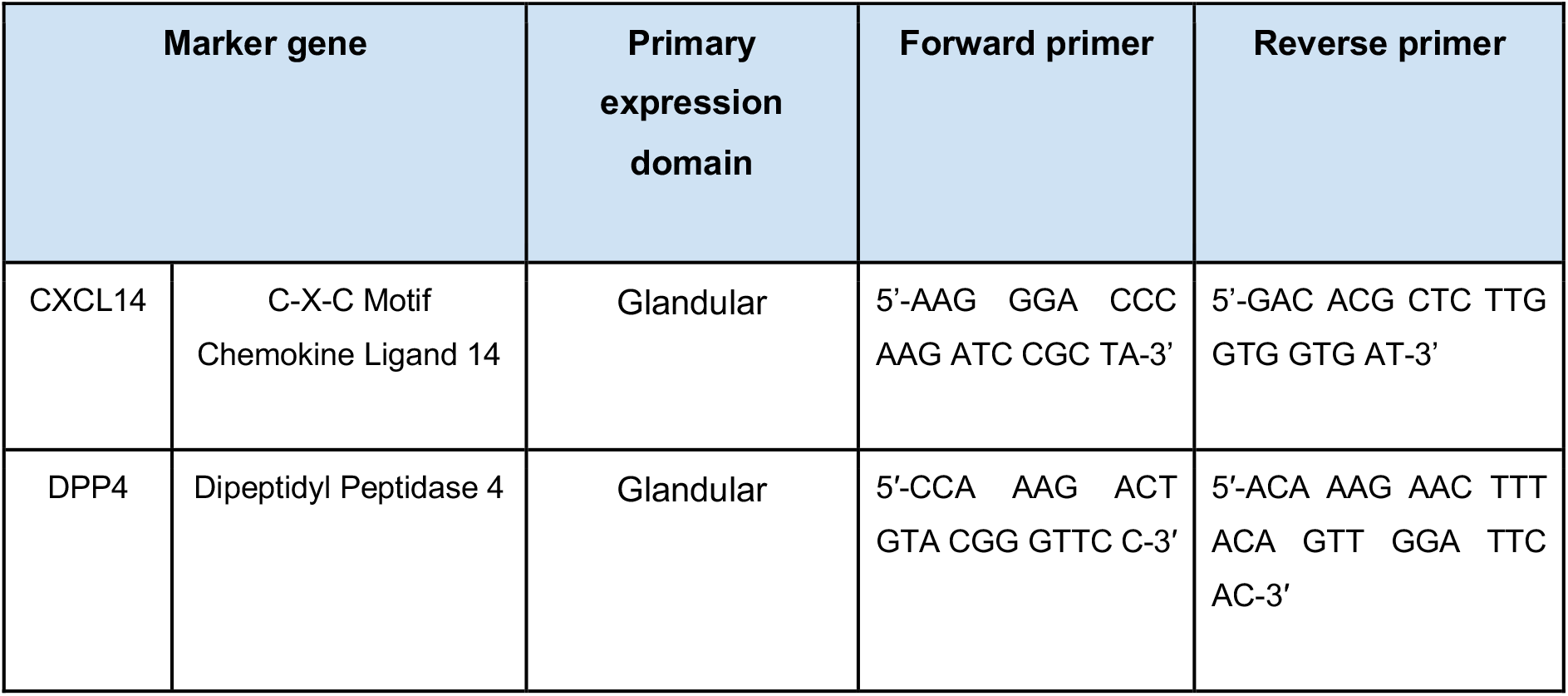
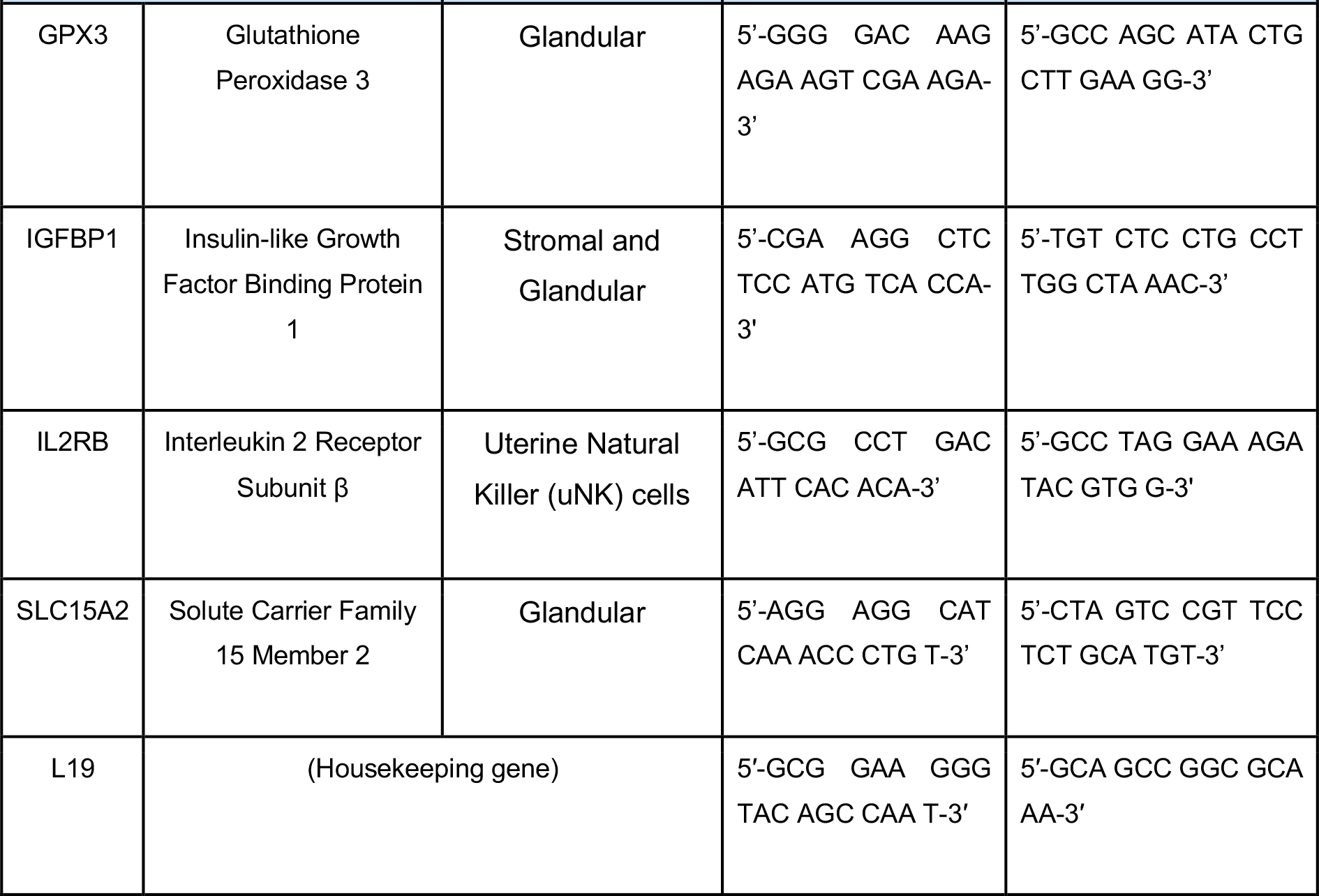
Timing marker genes used for the EndoTime method.

